# Integrated fragmentomic profile and 5-Hydroxymethylcytosine of capture-based low-pass sequencing data enables pan-cancer detection via cfDNA

**DOI:** 10.1101/2022.11.30.22282918

**Authors:** Zhidong Zhang, Xuenan Pi, Chang Gao, Jun Zhang, Lin Xia, Xiaoqin Yan, Xinlei Hu, Ziyue Yan, Shuxin Zhang, Ailin Wei, Yuer Guo, Jingfeng Liu, Ang Li, Xiaolong Liu, Wei Zhang, Yanhui Liu, Dan Xie

## Abstract

Using epigenetic markers and fragmentomics of cell-free DNA for cancer detection has been proven applicable. We further combine the two features and explore the diagnostic potential of the features on pan-cancer detection. We extracted cfDNA fragmentomic features from 191 whole-genome sequencing data and investigated them in 396 low-pass 5hmC sequencing data from four common cancer types and controls. We identified aberrant ultra-long fragments (220-500bp) of cancer samples in 5hmC sequencing data, both in size and coverage profile, and showed its dominant role in cancer prediction. Since cfDNA hydroxymethylation and fragmentomic markers can be detected simultaneously in low-pass 5hmC sequencing data, we built an integrated model including 63 features of both fragmentomic features and hydroxymethylation signatures for pan-cancer detection with high sensitivity and specificity (88.52% and 82.35%, respectively). We showed that fragmentomic information in 5hmC sequencing data is an ideal marker for cancer detection and that it shows high performance in low-pass sequencing data.

## Introduction

Displaying a trend of increase, more than 19.3 million individuals were diagnosed with cancer, and 10.0 million individuals died of cancer worldwide in 2020 (1). Currently, tissue biopsy is the most widely used method for cancer diagnosis and staging. However, owing to being an invasive examination method, tissue biopsy has difficulties in tracking clonal evolution, monitoring treatment response, and early detection of cancer and recurrence (2). Therefore, the need for the development of accurate liquid biopsy methods is urgent, with the potential to detect cancer and identify resistance mechanisms early, to quantify minimal residual disease, and to monitor treatment responses (2, 3).

Circulating cell-free tumor DNA (ctDNA) is a kind of tumor-derived material in the blood of patients with cancer and is mainly derived from cancer cells via apoptosis or necrosis (3, 4). ctDNA carries cancer-specific genetic mutations, epigenetic alterations, and fragmentomic aberrations, allowing for cancer detection and tissue-of-origin prediction in a non-invasive manner. Nevertheless, as cancer is a highly heterogeneous disease, biomarkers based on genetic and epigenetic aberrations often have limitations in practical applications. Recent studies have shown that cell-free DNA (cfDNA) fragmentation is a non-random process (5, 6), and it is thus possible to develop a generic approach for non-invasive cancer detection using cfDNA fragmentomics.

The fragmentomics analysis of cfDNA encompasses the sizes (7), coverage (7), nucleosome footprints (8), end-points (9, 10) and end-motifs (11) of cfDNA. For example, the size distribution of cfDNA fragments can indicate specific cellular biological changes (12, 13). Coverage in cfDNA sequencing can be used to analyze nucleosome positions (8, 14) and identify expressed genes (14). Fragmentation patterns of cfDNA can be characterized by deferentially phased fragment end signals, which were preferentially found in tissue-specific open chromatin regions (15). Practically, Jiang et al. showed that cfDNA fragments ending with particular genomic coordinates or motifs had higher degrees of association with hepatocellular carcinoma (HCC) (10, 11). Recently, the epigenetic marker 5-hydroxymethylcytosine (5hmC) has attracted attention in the field of cancer research due to its regulatory role in tissue-specific gene expression (16). The cfDNA captured by 5hmC comprises a subset of all circulating cfDNA and, therefore, may contain fragmentomic information. Integrative multi-omics analysis including the 5hmC profile of cfDNA improves the sensitivity and specificity of cancer detection, as demonstrated by recent studies (17, 18).

To highlight the potential of 5hmC sequencing data in cancer detection, we obtained a large cohort of cancer samples, including HCC, pancreatic ductal adenocarcinoma (PDAC), lung adenocarcinoma (LUAD), and glioblastoma (GBM). We then established a pan-cancer preferred end map based on WGS data from these four cancer types and checked their characteristics in low-pass 5hmC sequencing data. We hypothesized that the fragmentomic information would be consistent between WGS and 5hmC sequencing data and tested it by comparison of each feature. We built classification models and investigated the effect of combining 5hmC signatures with fragmentomic information in pan-cancer screening.

## Methods

### Sample collection and study design

In total, low-pass (mean mapped reads: 38.32 ± 8.02 million) 5hmC-seq data from 85 healthy controls and 311 cancer patients were included in this study. We obtained 5hmC sequencing data from 33 LUAD (18), 74 PDAC (37), 132 HCC (38), and 85 control (37) samples from three publications, and 72 GBM samples from an unpublished paper.

Plasma samples from 15 healthy controls and 59 cancer patients were collected and subjected to WGS (mean: 14×, range: 8.6-23.7×) to establish the preferred end marker set, while the low-pass WGS data from 19 healthy controls and 98 cancer patients (mean: 3×, range: 2.3-6.2×) were obtained to cross-check with low-pass 5hmC sequencing data. 13 of the samples with low-pass WGS sequencing had above 10x WGS sequencing data. The demographics and clinical characteristics of the cohort are summarized in Supplementary Tables 1 and 2. The design of the study is shown in Figure 1.

**Figure 1.**
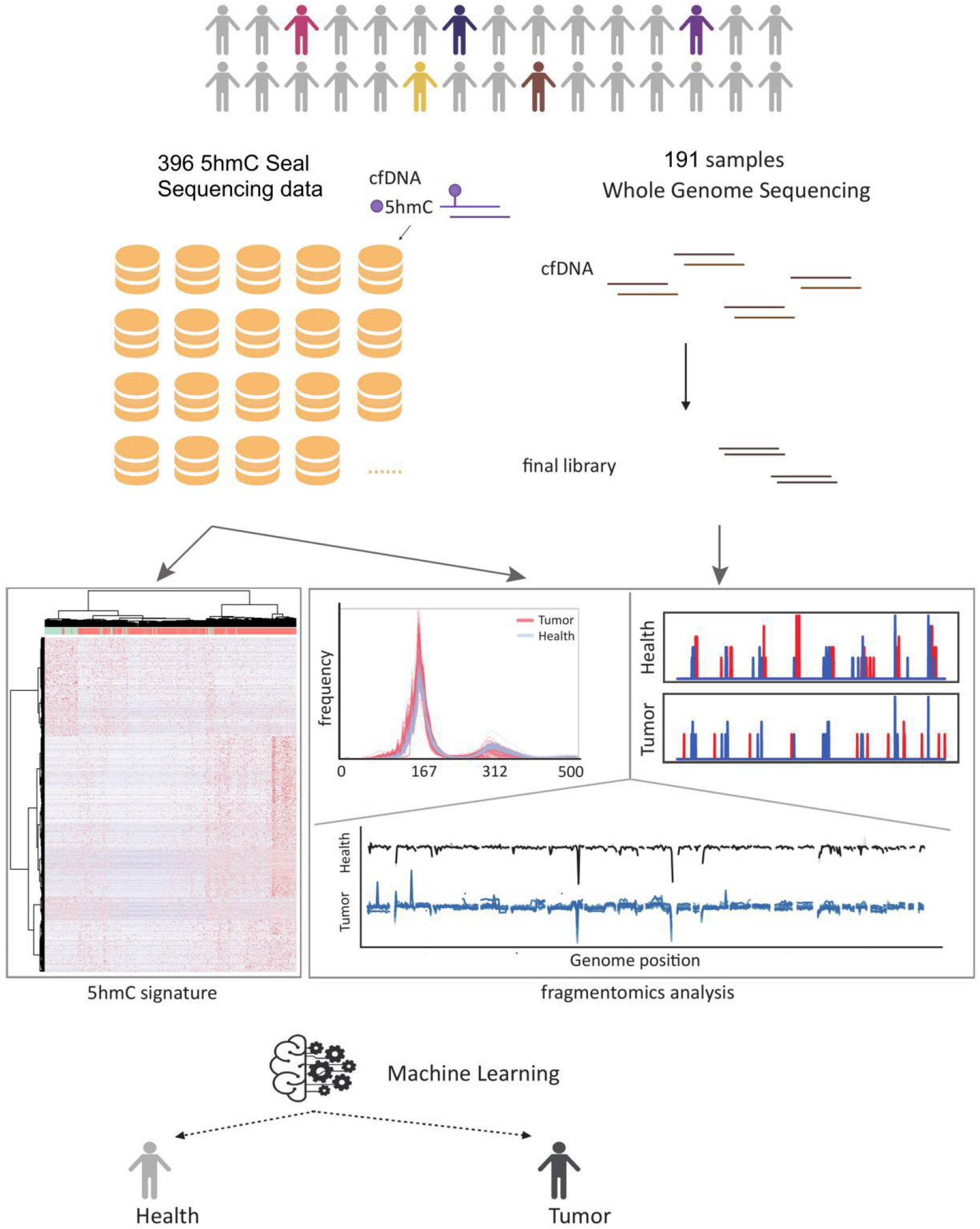
Overview of the study design. cfDNA from cancer patients and controls were sequenced with WGS. Available 5hmC sequencing data from the same four cancer types and controls were combined with WGS data to get the accurate fragmentomic information and upon it a machine learning model was built to distinguish healthy controls and cancer patients.

### Plasma sample collection and cfDNA WGS sequencing

We collected plasma samples from 46 GBM patients, 29 PDAC patients and 34 controls from West China Hospital. Plasma samples of 37 HCC patients and 32 LUAD patients were collected from Mengchao Hepatobiliary Hospital of Fujian Medical University and the First Affiliated Hospital of the Second Military Medical University, respectively. For every subject, the QIAseq cfDNA All-in-one Kit (Qiagen) was applied on the 2 ml plasma sample for the cfDNA extraction and library construction. Pair-end 150 bp sequencing of the libraries was performed on the Illumina Novaseq 6000 platform.

### cfDNA 5hmC profiling and sequencing

cfDNA extraction and 5hmC library construction was performed as previously described (28). Firstly, cfDNA was extracted from 2 ml plasma sample using QIAamp Circulating Nucleic Acid Kit (QIAGEN Inc., Valencia, CA, USA). Then, cfDNA (5–10 ng) ligated with sequencing adaptors was incubated in a 25 μL reaction solution containing HEPES buffer (50 mM, pH 8.0), 25 mM MgCl2, 60 μM N3-UDP-Glc (ActiveMotif, Carlsbad, CA, USA), and 12.5 U β-glucosyltransferase (NEB, Beverly, MA, USA) for 2 h at 37 °C. Next, 2.5 μL DBCO-PEG4-biotin (Sigma, Carlsbad, CA, USA) was directly added and incubated for 2 h at 37 °C. 10 μg sheared salmon sperm DNA (Life Technologies, USA) was added, then the Micro Bio-Spin 30 Column (Bio-Rad, Hercules, CA, USA) was used to purify the DNA following the instruction, and the final volume was adjusted to 25μL. After that, the purified DNA was incubated with 5 μL C1 streptavidin beads (Life Technologies, USA) in buffer 1 (5 mM Tris pH 7.5, 0.5 mM EDTA, 1 M NaCl and 0.2% Tween 20) for 30 min. The beads were subsequently undergone three 5-min washes each with buffer 1, buffer 2 (buffer 1 without NaCl), buffer 3 (buffer 1 with pH 9) and buffer 4 (buffer 3 without NaCl). Then, the beads were resuspended in water and amplified with 11 cycles of PCR amplification (initial denaturing at 98 °C for 45 s, followed by 11 cycles of denaturing at 98 °C for 15 s, annealing at 60 °C for 30 s, extension at 72 °C for 30 s, and a final extension at 17 °C for 5 min). The amplified products were purified using 0.8 × AMPure XP beads (Beck-man Coulter, Fullerton, CA, USA). Pair-end 150 bp sequencing was performed on the Illumina Novaseq 6000 platform.

### Fragmentomic profiling of cfDNA and cancer prediction analysis

The details of the preferred end, 5hmC signatures, size profile and coverage profile calculation are described in the Supplementary Material. We used a random forest model to distinguish healthy people from cancer patients using fragmentomic features. To estimate the prediction error, we used five cross-validations. Near-zero variance features were removed. The training, validation, and test sets account for 60%, 20%, and 20% of the data, respectively. The samples were selected randomly in a balanced way to keep the ratio of the number of cancer to non-cancer samples similar in the training, validation, and test subsets. Feature importance was calculated using the training data in each cross-validation run, and we sorted the features according to the mean value of feature importance. The number of features used in the final model was obtained by the highest AUC value in the validation set. To build an integrative model, we fitted the features selected by every feature-alone model into one random forest model, and performed feature selection, training, testing as described above. Random forest machine learning was implemented using the python package sklearn.ensemble.RandomForestClassifier with the following parameters: n_estimators = 100, criterion=“gini”.

### Statistics

All statistical analyses were performed using R version 3.6.1 and python 3.7.0. All tests were two-sided, and P values < 0.05 were considered statistically significant. The ROC-AUC plot and AUC value were implemented using sklearn.metrics.plot_roc_curve.

### Ethics

This study was approved by the Ethics Committee of Sichuan Cancer Hospital (SCCHEC-02-2016-005). The written informed consent was obtained from all participants.

## Results

### Constructing the cfDNA preferred end map

cfDNA fragments ending at specific genomic coordinates that were statistically more abundant than the Poisson distribution was an essential fragmentation signature referred to as the preferred end (10). We utilized WGS data from the four cancer types to build a preferred end map and checked the feature using low-pass data. The basis of utilizing cfDNA preferred end coordinates to indicate the occurrence and location of cancer was its ability to locate nucleosomes, the position of which is related to cell identity (19). To confirm this, we defined upstream (U) ends and downstream (D) ends by their genomic coordinate order on the reference genome calculated via alignment. We surveyed the distance distribution between adjacent U ends, D ends, and nucleosome centers, and the result suggested the preferred ends were enriched on the DNA linker (Supplementary Figure 1a-f). We further checked the distribution of the predicted nucleosome location based on non-malignant tissues at chr12: 34517269-34519122, and the preferred U and D ends of WGS data from controls (n=15) and HCC patients (n=15). As expected, the preferred U and D ends of the controls were enriched at the midpoint between two nucleosomes, which is the location of the DNA linker (Figure 2a), and U ends were found downstream of their nearest D ends. However, in HCC patients, U ends were upstream of their nearest D ends, and only D ends were found at one midpoint between two nucleosomes (Figure 2b), which indicated inconsistent nucleosome positions between HCC patients with controls. We observed the same abnormal preferred end distribution in GBM, LUAD and PDAC patients (Supplementary Figure 1g-i).

**Figure 2.**
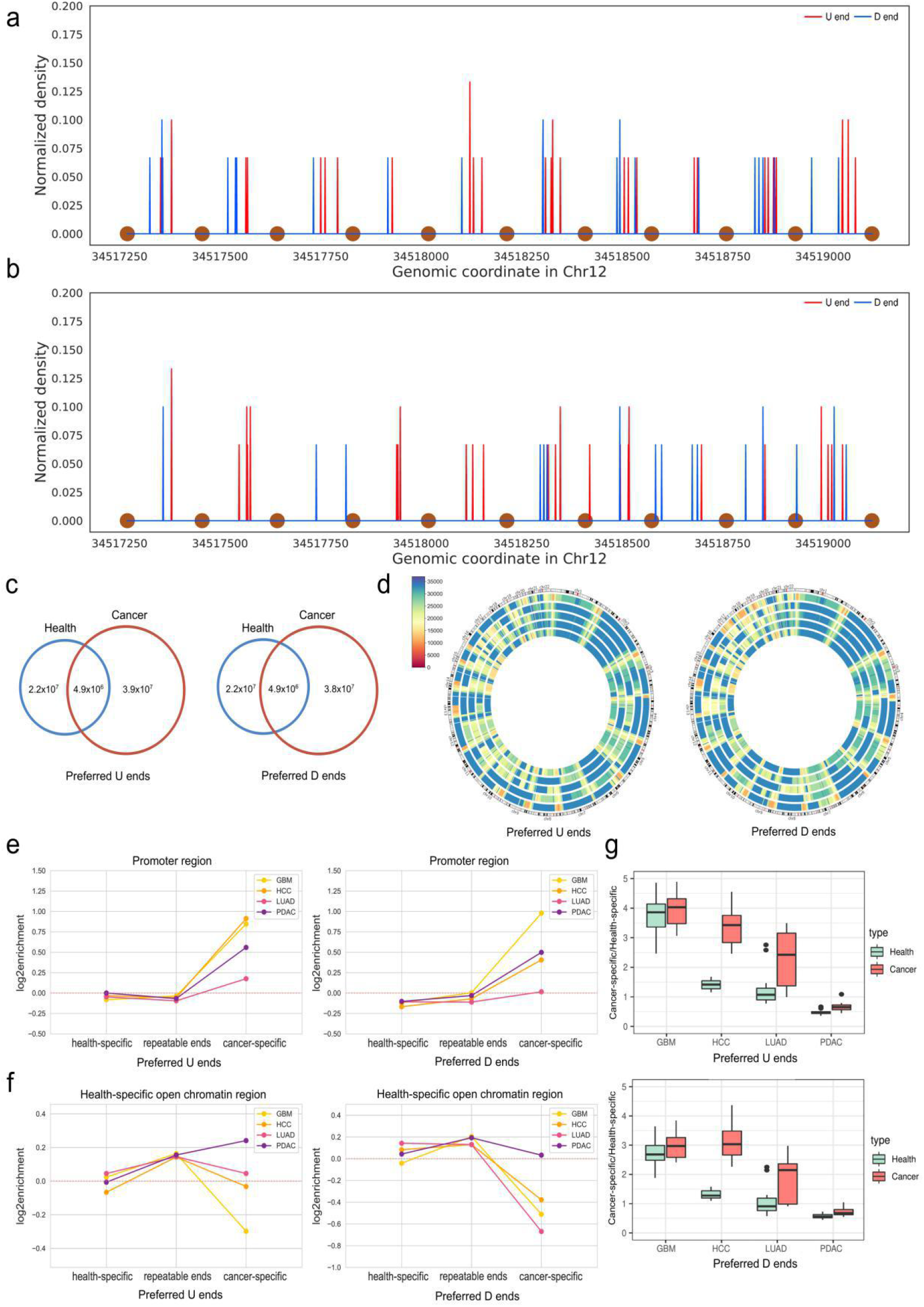
Genome-wide cfDNA fragment preferred end map construction. cfDNA preferred end signals in a nucleosome array region (chr12: 34517269-34519122) in healthy (a) and HCC (b) subjects. Brown dots at the bottom represent the predicted nucleosome center loci reported in a previous study (8, 22). (c) Venn diagram showing the intersection of the preferred U and D ends between the cancer cohort and the control cohort. (d) Circle diagram showing the density of preferred ends in 1 Mb windows in four types of cancer and in healthy controls (the order of sample sources from outer ring to inner ring are control, GBM, HCC, LUAD and PDAC). (e) Enrichment of preferred U/D ends in promoter regions. (f) Enrichment of preferred U/D ends in health-specific open chromatin regions. (g) Boxplots showing the ratios of the cancer-specific to health-specific preferred U/D ends in low-pass WGS data for the four cancer types (Wilcoxon rank-sum test).

By using WGS data, we constructed cfDNA fragment preferred end maps for the four cancer types and control (Figure 2c-d). Preferred end enrichment was observed in intron and open chromatin regions (Supplementary Figure 2). Additionally, we noticed that for every cancer type, both U and D preferred ends were more enriched in health-specific open chromatin regions than in cancer-specific open chromatin regions (Supplementary Figure 2). One explanation was the nucleosome pattern in the health-specific open chromatin regions was unstable, as the enrichment of both U and D preferred ends in healthy cfDNA were significantly lower than cancer patients (Mann‒Whitney, P<0.05). Another was the heterogeneous nature of cfDNA, as cfDNA can be derived from various tissue sources. To search for preferred ends specific to each of the four cancer types, we determined genome-wide cancer-specific preferred U/D end coordinates and health-specific preferred U/D end coordinates based on the Youden index. In total, we got 81,083 D and 80,505 U preferred ends. Intriguingly, compared with all repeatable ends (preferred ends appeared in at least two samples in each cancer type), cancer-specific preferred U/D ends were no longer enriched in health-specific open chromatin regions, but they showed enrichment in promoter regions and cancer-specific open chromatin regions (Figure 2e-f, Supplementary Figure 3-4). Furthermore, health-specific preferred U/D ends showed enrichment in intergenic regions and were not found in promoters, as cancer-specific preferred ends were (Supplementary Figure 4). To cross check the cancer specificity of the preferred ends in low-pass WGS data, we calculated a ratio, which was the ratio of detected cancer-specific preferred end numbers to health-specific preferred end numbers for each sample. The results showed significantly higher ratios of cancer-specific to health-specific preferred U/D ends in the three cancer types (except GBM) than in the control (Figure 2g, Mann‒Whitney, p value<0.001). In addition to the difference between cancer and health, statistically significant differences in the ratios between cancer types were observed (Supplementary Figure 5).

### Fragmentomic features in 5hmC data

We checked the batch effect of the 5hmC sequencing data based on size profile and coverage profile, and found no clustering of samples according to cancer types (Supplementary Figure 6).

The cfDNA captured by 5hmC marks comprised a subset of DNA in whole plasma. In theory, one sequenced fragment should contain at least one 5hmC. Hence, we investigated the consistency of the size profile between WGS data and 5hmC sequencing data. According to the results, fragment sizes of 5hmC cfDNA in the four cancer samples were significantly shorter than those in the control (Mann‒Whitney test, p value<0.001, pan-cancer Median N50 = 174 bp, control Median N50 = 210 bp), though the difference between cancer and control groups in WGS data was only observed for HCC and LUAD (Figure 3a). Next, we confirmed that the cfDNA size profile of the 5hmC sequencing data was consistent with that of the WGS data in the size range of 0-220 bp (Figure 3b), with both data types displaying a dominant peak at 167 bp (Supplementary Figure 7a). A difference between the cancer and control samples in both short (100-150 bp) and long (151-220 bp) cfDNA fragments (7) was found in the 5hmC data, as in the WGS data (Supplementary Figure 7b-c), though the 5hmC data showed a unique secondary peak at ∼320 bp (Figure 3c, Supplementary Figure 7a). In the WGS data, cfDNA fragments ranging from 0-220 bp accounted for nearly 100% of fragments ranging from 0-800 bp, but the percent value of the same size interval in the 5hmC sequencing data ranged from 31.8% to 99.6% (Figure 3f). Thus, we confirmed the presence of ultra-long fragments (221-500 bp) in the 5hmC cfDNA. We suspected that the existence of ultra-long fragments may have resulted from the capture-based technique applied in 5hmC sequencing, and we consequently assessed the relationship between fragment length and 5hmC peak number. In ultra-long fragments, the percentage of fragments with more than one peak was significantly higher than that in non-ultra-long fragments (0-220 bp) (Mann‒Whitney test, p value<0.001) (Figure 3d). Indeed, enrichment analysis of the ultra-long fragments with more than one peak revealed primary enrichment at CpG islands (Supplementary Figure 7d). This indicated that the capture-based sequencing technique is more likely than WGS to capture ultra-long cfDNA fragments. Another finding was that in all cancer types, the percentage of ultra-long fragments was significantly lower than that in the control (Supplementary Figure 7b). We examined the percentage of ultra-long fragments with more than one 5hmC peak in the four cancer types and the control and found that the value was significantly lower in the former (Figure 3e). As reported by a recent study, the stability of circulating DNA derives mostly from the nucleosome structure (21). We hence evaluated the proportion of 5hmC cfDNA fragments at 146 bp, 166 bp, 312 bp, and 332 bp, which indicates the length of one mono-nucleosome and one di-nucleosome (plus the linker size). The result showed a higher proportion of mono-nucleosome-sized cfDNA fragments and a lower proportion of di-nucleosome-sized cfDNA fragments in the cancer group than in the control (Supplementary Figure 7e).

**Figure 3.**
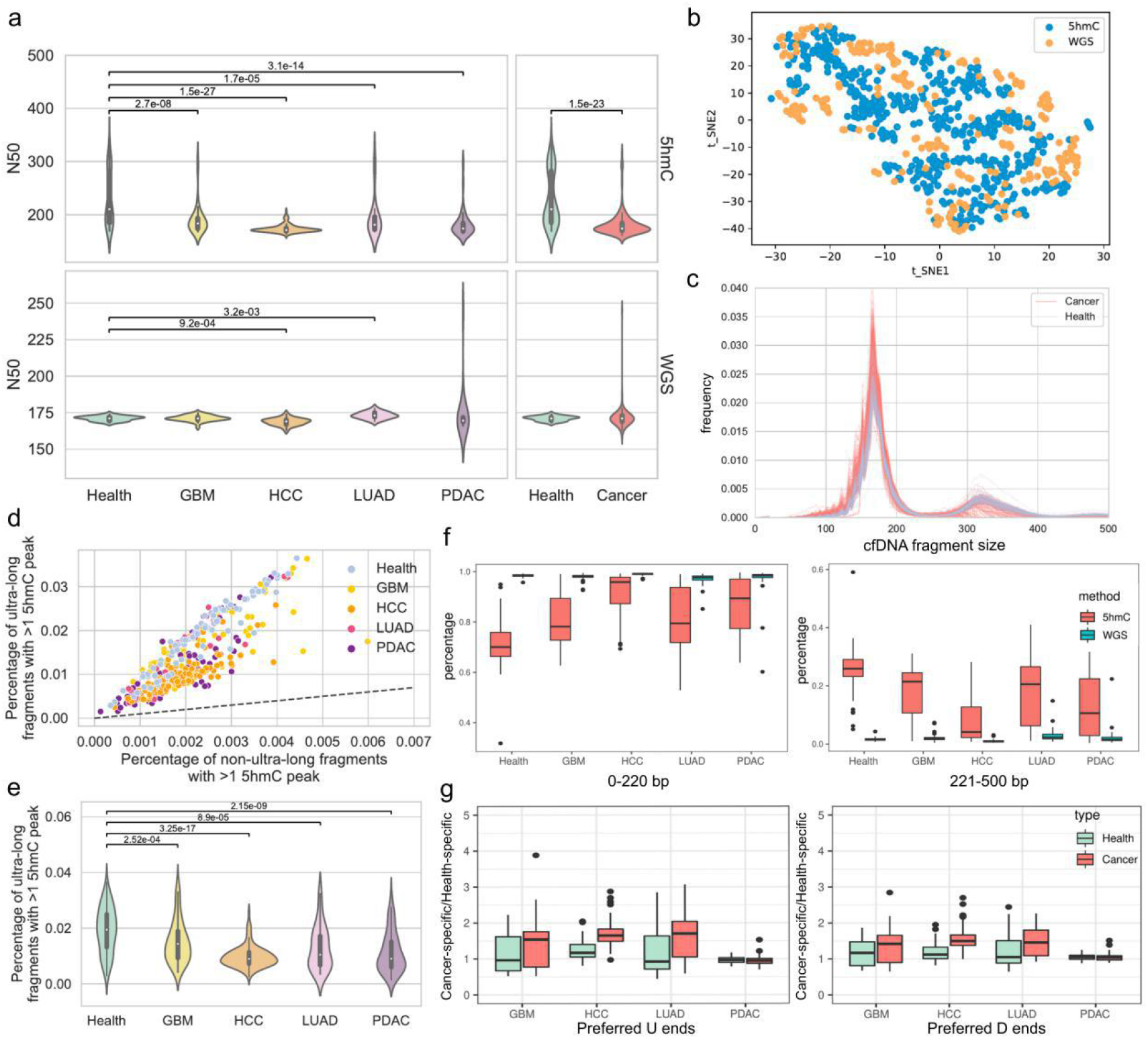
cfDNA fragmentomic information in 5hmC sequencing data. (a) Violin plots showing fragment size profile comparison between cancer samples and controls in terms of N50 (Mann‒Whitney test). (b) tSNE plot showing 5hmC sequencing data and WGS data by fragments below 220 bp. (c) The cfDNA size profile of 5hmC sequencing data. (d) Comparison of the percentage of fragments with more than one 5hmC peak between ultra-long and non-ultra-long fragments in every sample. The dashed line indicates the diagonal line. (e) Violin plots showing the percentage of ultra-long fragments with more than one 5hmC peak in cancer samples and controls (Mann‒Whitney test). (f) Boxplots showing the percentage of fragments ranging from 0-220 bp and 221-500 bp among fragments ranging from 0-800 bp in WGS data and 5hmC sequencing data (Wilcoxon rank-sum test). (g) Boxplots showing the ratios of the cancer-specific to health-specific preferred U/D ends in each of the four cancer types (Wilcoxon rank-sum test).

We also explored the cancer specificity of the pan-cancer preferred end set built upon WGS data in the 5hmC sequencing data by calculating the ratio of cancer-specific to health-specific preferred U/D ends. The ratios were significantly increased in cfDNA samples in three cancer types (except PDAC) compared with healthy individuals (Figure 3g, Supplementary Figure 5c-d, Mann‒Whitney, p value<0.005). Coverage profiles in 5hmC sequencing data also showed a higher variance than WGS data in samples from control and cancer (Supplementary Figure 8). We explored the coverage patterns of short, long, and ultra-long cfDNA fragments at the 5 Mb bins of the genome. Similar to WGS data (Supplementary Material), cancer patients exhibited multiple unstable genomic regions: coverage of short/long/ultra-long fragments was inconsistent in cancer patients but consistent in controls (Figure 4a, Supplementary Figure 9a). We analyzed the genome-wide coverage profile correlation of cancer patients to controls in the 5hmC data and found that the coverage profiles were consistent among controls and that the correlation value in cancer patients was significantly lower (Wilcoxon rank-sum test, p value<0.005). Importantly, the largest difference between cancer samples and controls was found with respect to the coverage profile of ultra-long fragments (Wilcoxon rank-sum test, p value<0.005) (Figure 4b).

### Genomic distribution of 5hmC in cancer and control cohorts

To gain an understanding of the genomic regions associated with hydroxymethylation in cfDNA, we first determined 5hmC-enriched loci in the control and four types of cancer, which were detected as peaks via MACS2 (22). 5hmC-modified regions among samples were compared in 1 kb bins on the reference genome, and thus a consensus list of the absence and presence of 5hmC-modified peaks among the samples was obtained. 5hmC signatures were enriched over genic features, most significantly in the promoter, 5’UTRs, 3’UTRs, exons, and transcription end sites (TESs) (Figure 5a). Comparison of 5hmC signature enrichment in the four types of cancers with the control revealed significant differences, whereby 5hmC peak enrichment was lower in all cancer types than in the control over promoters, 5’UTRs, exons, and CpG islands (Wilcoxon rank-sum test, p value<0.001) (Figure 5a).

Differential analysis of 5hmC-modified peaks between cancer patients and controls by Fisher’s exact test detected 1,010, 6,395 and 773 differentially modified 5hmC peaks in GBM, HCC and PDAC, respectively. Moreover, this pan-cancer 5hmC peak set was able to separate most cancer samples from the control (Figure 5b). KEGG enrichment analysis of genes located within the differentially modified 5hmC peaks revealed associated oncogenesis pathways for each cancer type (Supplementary Figure 10), such as the neurotrophin signaling pathway and platelet activation for GBM (23, 24), the MAPK signaling pathway and focal adhesion for HCC (25, 26), and the FoxO signaling pathway and insulin signaling pathway for PDAC (27).

### Cancer detection by combining fragmentomic features and 5hmC signatures using 5hmC sequencing data

As 5hmC sequencing data retain both 5hmC signatures and fragmentomic information, it is expected to theoretically show high sensitivity and specificity in cancer detection. A receiver operator characteristic (ROC) curve was used to evaluate the performance of the classifier. In total, 53 5hmC signatures were selected, and the AUC of the classifier was 0.876 in the validation set (sensitivity = 82.26%, specificity = 82.35%) and 0.872 in the test set (sensitivity = 81.97%, specificity = 82.35%) (Figure 5d-e, Supplementary Figure 11 c).

Overall, the fragmentomic information in the 5hmC sequencing data performed well. Using the size profile as features, 40 were selected to build a pan-cancer prediction model, among which 32 features were of the ultra-long and longer size range (>= 220 bp) (Figure 5c). The AUC value was 0.981 in the validation set (sensitivity = 93.55%, specificity = 94.12%) and 0.882 in the test set (sensitivity = 71.43%, specificity = 88.24%) (Figure 5d-e). Using preferred ends as features, 37 were selected to build a pan-cancer prediction model. ROC analysis showed an AUC value of 0.940 in the validation set (sensitivity = 83.87%, specificity = 94.12%) and 0.899 in the test set (sensitivity = 73.02%, specificity = 94.12%) (Figure 5d-e, Supplementary Figure 11a). Regarding the coverage profile of the 5hmC sequencing data, we explored the genomic distribution of short, long, and ultra-long cfDNA fragments on a 100 kb window of the genome. This model achieved an AUC value of 0.946 in the validation set (sensitivity = 80.65%, specificity = 94.12%) and 0.882 in the test set (sensitivity = 71.43%, specificity = 88.24%) (Figure 5d-e).

Finally, we constructed an integrated cancer screening model by combining fragmentomic features and 5hmC signatures in 5hmC data. Sixty-three features, including 10 5hmC signatures, 24 size profile features, 21 coverage profile features, and 8 preferred end features, were selected to build the pan-cancer prediction model. The AUC value was 0.927 in the validation set (sensitivity = 93.44%, specificity = 88.24%) and 0.920 in the test set (sensitivity = 88.52%, specificity = 82.35%) (Figure 5d-e). The performance of single-cancer detection is depicted in Supplementary Figure 11b. With data from the four cancer types, we further explored tissue-of-origin prediction. The performance of the random forest model with 5hmC fragmentomic features and the integrated model are shown in Supplementary Tables 3-5.

## Discussion

cfDNA 5hmC signatures are reported to have potential for cancer detection, such as in lung cancer (28), hepatocellular carcinoma (28), colon cancer (29), gastric cancer (29), and pancreatic cancer (17, 30). The large-scale cohort of 5hmC sequencing data utilized in this study additionally shows the potential of cfDNA 5hmC signatures for cancer detection in glioblastoma and pan-cancer. Besides, we further examined the classification effect using fragmentomic features in tissue-of-origin prediction of cancer. Most importantly, we explored cfDNA fragmentomic information in 5hmC sequencing data and found that it is possible to detect cfDNA hydroxymethylation and fragmentomic markers simultaneously. The size profile, preferred end, and coverage profile in 5hmC sequencing data showed a large difference between cancer patients and healthy individuals. The integrated model covering the 5hmC signature, size profile, preferred end, and coverage profile contained more information than the model with the 5hmC signature alone, as revealed by higher sensitivity and specificity in pan-cancer detection.

Previous work reported a difference in the size profile between specific cancer patients or pan-cancer patients and healthy controls (12, 13, 31), though the analyses only focused on lengths less than 320 bp. Here, we characterized ultra-long fragments in 5hmC sequencing data and identified their size and coverage profile aberrations in cancer samples. Although it is commonly acknowledged that ctDNA tends to be more fragmented than cfDNA in normal tissue (12, 31), we found an even larger size difference in cfDNA between cancer samples and controls in 5hmC data owing to ultra-long fragments. Hence, we further expanded the size range to 800 bp by setting 80 adjacent, non-overlapping bins to capture 10-bp interval signals (32). The coverage profile is another major improvement we made with regard to previously reported models based on fragmentation information (9). Indeed, only the ratio of short to long cfDNA fragments has been analyzed in previously reported models, with information on cfDNA ultra-long fragments being lost. Our results suggest that the coverage distribution of size-selected cfDNA fragments in ultra-long fragments has the strongest inconsistency between control and cancer samples (Figure 4a). Of note, the classification models selected the majority of features from ultra-long and longer size range fragments in the coverage profile model (34/42), size profile model (32/40) (Figure 5c, Supplementary Figure 12c) and integrated model (41/63), suggesting that ultra-long features are a major contribution to the classification model. Further research on the mechanism of the aberration of the captured ultra-long fragments in cfDNA from cancer samples may provide insights into the fundamental biological properties of plasma cfDNA from patients with cancer.

**Figure 4.**
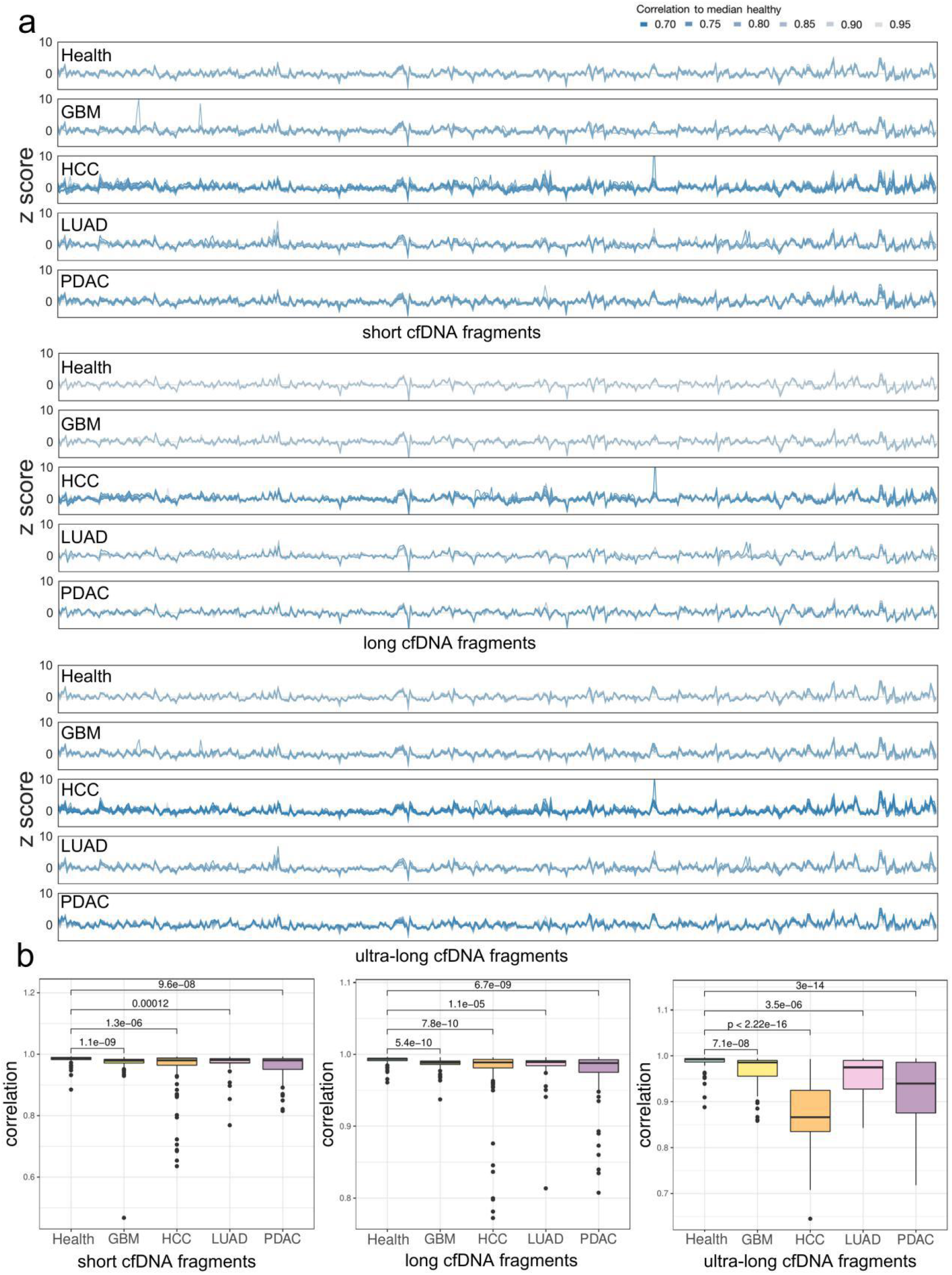
Size-selected coverage profile and correlation to median health of cfDNA fragments in 5hmC sequencing data. (a) Genome-wide coverage profile of short, long, and ultra-long cfDNA fragments; color indicates sample-wise correlation to median health. (b) Coverage profile correlation to median healthy in control and four types of cancer in short, long, and ultra-long cfNDA fragments (Wilcoxon rank-sum test, p value<0.005).

**Figure 5.**
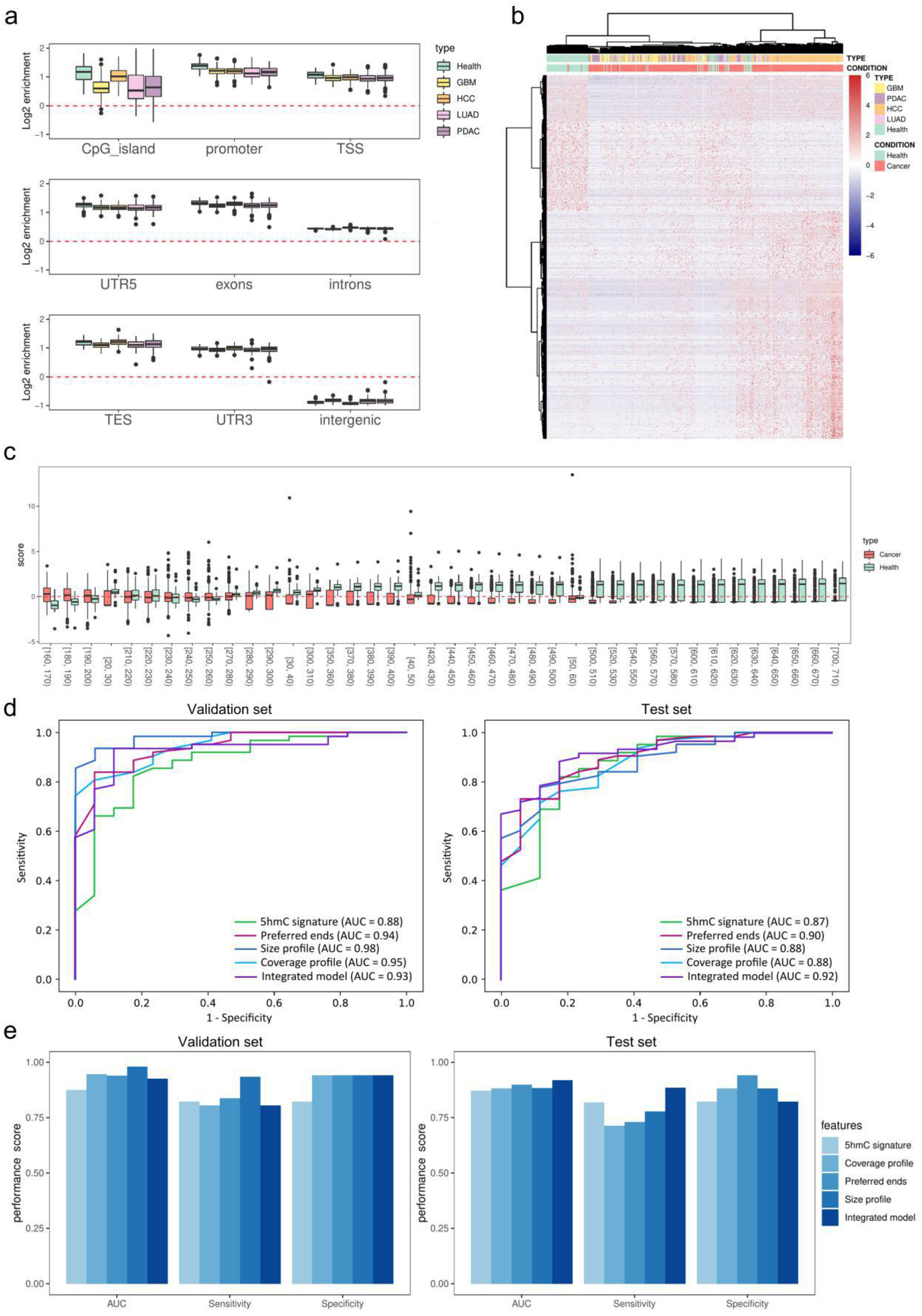
Performance of cancer detection combining fragmentomic features and 5hmC signatures using 5hmC sequencing data. (a) Enrichment of 5hmC in genomic features in control and four types of cancer (Wilcoxon rank-sum test, p value<0.001). (b) Heatmap of differentially modified 5hmC peaks. (c) Selected size profile features in the size profile model. (d) ROC curves for the validation set and test set in pan-cancer diagnosis. (e) Performance evaluation of validation set and test set in pan-cancer diagnosis.

In our work, we constructed a single-base resolution preferred end set based on a set of WGS data. Our results indicated that the preferred end set had good generalization ability because the differences between patients with cancer and controls were also shown in other independent data sets, e.g., low-pass WGS data and 5hmC data. It has been reported that referred end coordinates differ between HCC patients and healthy people at a whole-genome scale (10). Apart from HCC, our results suggest the cfDNA preferred end can also be used to distinguish patients with GBM, LUAD, and PDAC. We herein discussed the distribution of cfDNA preferred ends in genomic features, open chromatin regions, and nucleosome structure. The distribution of cfDNA preferred ends can reflect nucleosome positioning, though there is no apparent preference in genomic features and open chromatin regions, consistent with previous work (15). However, informative cancer-specific preferred ends were found to be significantly enriched in promoter and far away from health-specific open chromatin regions (Figure 2e-f, Supplementary Figure 4c-d, Supplementary Figure 5c-d).

In summary, our results indicate that cfDNA fragmentomic analysis with a nonlinear classification algorithm using low-pass 5hmC sequencing data may provide a simple, low-cost, and highly effective method for cancer detection. The success of utilizing ultra-long fragments in 5hmC sequencing data as biomarkers indicates the potential of other capture-based sequencing approaches, such as cell-free methylated DNA immunoprecipitation and high-throughput sequencing (cfMeDIP–seq) (33), in the diagnosis of cancer. Notably, third-generation sequencing methods are expected to detect long cfDNA molecules, which have been utilized in non-invasive prenatal testing (34). As third-generation sequencing methods can detect sequence context and epigenetic modification at the same time, they can provide simultaneous analysis of genome-wide genetic, fragmentomic and epigenetic detection (35, 36). Moreover, standardized and effective workflows for analysis of cfDNA fragments need to be developed.

## Data availability

The 5hmC sequencing data for controls, LUAD, HCC and PDAC were publicly available as described in the Method section. The 5hmC sequencing data for GBM can be accessed at https://ngdc.cncb.ac.cn/gsa-human/s/15mfS98o.

## Acknowledgments

This study was supported by the 1·3·5 project for disciplines of excellence, West China Hospital, Sichuan University (ZYYC20006) to D. Xie; Thousand Talents Program of the West China Hospital (0040205401F58) to D. Xie; Sichuan Provincial Foundation of Science and Technology (2020YFS0051) to D. Xie, (2017SZ0006) to Y. Liu; Clinical Research Innovation Project, West China Hospital, Sichuan University (19HXCX009) to Y. Liu; the San Hang Program of the Second Military Medical University, Medical basic research project of the First Affiliated Hospital, the Second Military Medical University (2021JCMS16) to W. Zhang; the Science and technology project of Sichuan Province (2021YFS0109) to A. Li; Post-Doctor Research Project, West China Hospital, Sichuan University (20HXBH035) to S. Zhang; the high quality development of Guang ’an People’s Hospital (21FZ003) to A. Wei.

## Author Contributions

### Conception and design

Dan Xie, Zhidong Zhang, Xuenan Pi and Lin Xia.

### Collection and assembly of data

Jun Zhang, Xiaoqin Yan, Shuxing Zhang, Ailin Wei, Jingfeng Liu, Ang Li, Xiaolong Liu, Wei Zhang and Yanhui Liu. Data analysis and interpretation: Zhidong Zhang, Xuenan Pi, Chang Gao, Xinlei Hu, Xiyue Yan and Yuer Guo. Manuscript writing: Zhidong Zhang and Xuenan Pi. All authors read and approved the final version of the manuscript.

