## Supplementary Material for "Integrated fragmentomic profile and 5-Hydroxymethylcytosine of capture-based low-pass sequencing data enables pan-cancer detection via cfDNA"

### Methods

#### Data processing

Primary processing of whole-genome NGS data for cfDNA samples was performed using the Illumina BaseSpace Sequence Hub to generate sample specific FASTQ output. Cutadapt (1) was used to trim adaptor sequences of reads. The remaining reads were aligned to the human reference genome (UCSC, version hg19) using BWA-MEN read alignments. Read pairs with a MAPQ score below 20 for either read and PCR duplicates were removed via Samtools (2).

#### Identification of the open chromatin regions

The four types of cancer-associated open chromatin regions used in our study were built using ATAC-seq data reported in a previous study (3). The peak calling, iterative removal procedure, and quality control process were adopted as described (3). We downloaded the "cancer type-specific peak set", which is a set of high-quality, reproducible, fixed-width peaks for each of the cancer types. Then, the health-associated open chromatin regions was obtained from the ATAC-seq data reported in a previous study (4). In our study, we defined the health-associated open chromatin regions as the open chromatin regions obtained from peripheral blood mononuclear cells. Finally, four types of cancer-specific open chromatin regions were constructed by subtracting the peaks in health-associated open chromatin regions using Bedtools (5), and a list of health-specific open chromatin regions was constructed by subtracting the peaks in any type of cancer-associated open chromatin regions using Bedtools (5).

**Fragmentomic analysis of WGS data and 5hmC sequencing data from cfDNA**

The size profile was analyzed by calculating the proportion of cfDNA with a length of 11-20bp, 21-30bp, 31-40bp, 41-50bp, … 791-800bp. The 10bp window width was set because it had a better classification effect than the window width of 1bp or 5bp. Autosomes in Hg19 were tiled into 28720 adjacent, non-overlapping 100 kb bins. cfDNA fragments that fell in the Duke blacklisted regions (http://hgdownload.cse.ucsc.edu/goldenpath/hg19/encodeDCC/wgEncodeMapability/) were excluded. Short fragments were defined as between 100 to 150 bp in length, long fragments were defined as between 151 to 220 bp in length, and ultra-long fragments were defined as ranging from 221 to 500 bp, which was only analyzed in 5hmC sequencing data. To account for GC effects on coverage in plasma, we applied locally weighted scatterplot smoothing (LOWESS) regression analysis as previously described (6, 7). In detail, we set a span of 0.75 to the scatter plot of the average fragment GC versus fragment coverage in every 100 kb bin. This LOWESS regression analysis was performed separately for short, long, and ultra-long fragments in order to account for possible differences in coverage due to GC content. We obtained residuals that were uncorrelated with GC content by subtracting the predictions explained by GC content from the LOWESS model. Adding back the genome-wide median estimates of coverage, the residuals were returned to the original scale. Every sample was scaled to have mean zero and unit standard deviation to eliminate deviations caused by sequencing depth. This process was repeated for each sample.

The definition of U and D preferred end coordinates were the same as previously reported (8, 9), which was used to evaluate the frequency of nuclease cleaved at a locus on the genome.

Firstly, the actual value of cfDNA fragments ends at a certain locus was calculated by mapping cfDNA fragments to the hg19 reference genome. Based on the assumption that all cfDNA fragments were 166bp in length if the fragmentation was random, the two ends (U and D) of each fragment would be evenly distributed across a region 166bp upstream and downstream of the locus. A Bonferroni p-value was calculated to determine if a particular locus was an end that was significantly overrepresented based on a Poisson distribution. This test was performed separately for U and D ends.

1) $\mathrm{N_{actual}}$ : actual number of molecules terminating at a particular nucleotide

2) $\mathrm{N_{predict}} = \frac{\text{Coverage numebr}}{166}$

3) $\mathrm{P-value} = \mathrm{Poisson}(\mathrm{N_{actual}}, \mathrm{N_{predict}})$

A Bonferroni p-value of < 0.01 was considered as a cutoff to define the preferred ends.

**cfDNA 5hmC peak calling**

5hmC peak calling was carried out using MACS2 (10) with parameters "-f BED -t {pebed} --nolambda --nomodel --extsize 166 -g hs" to identify candidate 5hmC modified regions. The peak summits defined by MACS2 were extended by 250bp on a fixed width of 501bp, which were defined as 5hmC modified regions. All 5hmC modified regions were filtered by the Duke blacklisted regions (http://hgdownload.cse.ucsc.edu/goldenpath/hg19/encodeDCC/wgEncodeMapability/), and peaks on chromosomes X, Y or mitochondrial genome using bedtools (5). Then, we normalized MACS2 peak score (-log10(p-value)) as "score per million" which was calculated by dividing each peak score by the sum of all of the peak scores in the given sample divided by

1 million in order to reduce the MACS2 score deviation caused by read depth or quality. Peaks with score per million >= 5 would be selected as final 5hmC modified regions.

**Type-specific preferred ends calculation**

We constructed genome-wide cancer-specific preferred U/D end coordinates and health-specific preferred U/D end coordinates in our WGS data. For every genomic locus, we firstly calculated the percentage of the preferred end occurrences among all samples from one type (Health, PDAC, LUAD, HCC, GBM). Then, we calculated the Youden index for every type of cancer vs. controls. The definition of Youden index is sensitivity+specificity-1.

$$\mathrm{y} = \mathrm{p}_{\mathrm{cancer}} + (1 - \mathrm{p}_{health}) - 1$$

$$= \mathrm{p}_{\mathrm{cancer}} - \mathrm{p}_{health}$$

$\mathrm{y}$ stands for Youden index, and $\mathrm{p}$ stands for occurrences percentage of the preferred end from one type. Finally, we determined the cancer-specific and health-specific preferred ends based on the Youden index with a value above 0.4.

**Prediction model for tumor tissue-of-origin classification**

For cancer patients, a random forest model was trained to classify the tissue of origin using tissue-of-origin features in 5hmC signature, size profile, preferred end coordinates, or coverage profile. Similarly, to estimate the prediction error, we used five cross-validations. Near-zero variance features were removed. The training, validation, and test set account for 60%, 20%, and 20% of the data, respectively. The samples were selected at random in a balanced way to keep the proportions of four types of cancer samples similar in both the training, validation, and test subsets. In the training set, five-fold cross-validation was used to rank the features according to their importance. Feature importance was calculated on the training data in each cross-validation run and we sorted the features according to the mean value of feature importance. In the validation set, we selected the set of features with the highest AUC and constructed the classifier. Random forest machine learning was implemented using the python package sklearn.ensemble.RandomForestClassifier with parameters: n_estimators = 50 , criterion="gini". The predicted class of an input sample is a vote by the trees in the forest, weighted by their probability estimates.

#### Results

##### Size and coverage profile of cfDNA in WGS data

The WGS data showed that cancer patients' size profile was more variable than healthy controls (Supplemental Figure 9.B). We found that healthy individuals had consistent cfDNA long/short fragments coverage profiles at the 5 Mb bins of the genome. In contrast, cancer patients had various unstable genomic regions, where inconsistency of the short/long fragments coverage profiles between individuals was observed (Supplemental Figure 9A). We further performed a genome-wide correlation analysis of the size-selected cfDNA fragments coverage profiles, by comparing the coverage profile of each sample to the median coverage profile of healthy controls. We found that the coverage profiles were consistent among healthy controls, whereas the correlation value in cancer patients was significantly lower ($P<0.005$, Wilcoxon rank-sum test) (Supplemental Figure 9C).

**Supplemental Table 1. Demographics information of the subjects with 5hmC sequencing**

| | GBM (n=72) | HCC (n=132) | PDAC (n=74) | LUAD (n=33) | Control (n=85) |
|---|---|---|---|---|---|
| Age, average±SD | 50.11±13.66 | 57.62±11.24 | 62.48±10.48 | 59.87±9.12 | 51.93±12.50 |
| Gender, n | | | | | |
| Male | 37 | 106 | 33 | 17 | 27 |
| Female | 35 | 26 | 29 | 15 | 51 |
| NA | | | 22 | 1 | 7 |
| TNM, n | | | | | |
| I | | 56 | 11 | 1 | |
| II | 28 | 47 | 10 | | |
| III | 9 | 29 | 20 | 9 | |
| IV | 33 | | 19 | 22 | |
| NA | 2 | | 14 | 2 | |

**Supplemental Table 2. Demographics information of the subjects with WGS**

| | GBM (n=46) | HCC (n=37) | PDAC (n=29) | LUAD (n=32) | Control (n=34) |
|---|---|---|---|---|---|
| Age, average±SD | 55.05±14.16 | 57.72±10.27 | 59.4±11.49 | 62.33±8.79 | 57.09±9.98 |
| Gender, n | | | | | |
| Male | 26 | 27 | 21 | 15 | 9 |
| Female | 11 | 9 | 6 | 9 | 23 |
| NA | 9 | 1 | 2 | 8 | 2 |
| TNM, n | | | | | |
| I | | 19 | 2 | | |
| II | 1 | 12 | 7 | 3 | |
| III | 2 | 5 | 3 | 8 | |
| IV | 34 | | 6 | 10 | |
| NA | 9 | 1 | 11 | 11 | |

**Supplemental Table 3. Identification of tissue-of-origin by 5hmC signatures for cancer patients (Features number: 41)**

| Cancer | Validation set | | | | Test set | | | |
|---|---|---|---|---|---|---|---|---|
| type | Precision | Recall | F1-socre | Patients | Precision | Recall | F1-socre | Patients |
| GBM | 0.67 | 0.57 | 0.62 | 14 | 0.73 | 0.62 | 0.67 | 13 |
| HCC | 0.67 | 0.81 | 0.73 | 27 | 0.72 | 0.81 | 0.76 | 26 |
| LUAD | 0.33 | 0.17 | 0.22 | 6 | 0.20 | 0.14 | 0.17 | 7 |
| PDAC | 0.64 | 0.69 | 0.62 | 15 | 0.50 | 0.53 | 0.52 | 15 |
| All types | Accuracy：64.5%；F1-socre: 0.548 | | | | Accuracy: 62.3%；F1-score: 0.528 | | | |

**Supplemental Table 4. Identification of tissue-of-origin by preferred ends for cancer patients in 5hmC sequencing data (Feature number: 42)**

| Cancer | Validation set | | | | Test set | | | |
|---|---|---|---|---|---|---|---|---|
| type | Precision | Recall | F1-socre | Patients | Precision | Recall | F1-socre | Patients |
| GBM | 0.82 | 0.60 | 0.69 | 15 | 0.64 | 0.50 | 0.56 | 14 |
| HCC | 0.62 | 0.92 | 0.74 | 26 | 0.60 | 0.96 | 0.74 | 27 |
| LUAD | 0.67 | 0.33 | 0.44 | 6 | 1.00 | 0.29 | 0.44 | 7 |
| PDAC | 0.78 | 0.47 | 0.58 | 15 | 0.57 | 0.27 | 0.36 | 15 |
| All types | Accuracy：67.7%；F1-socre: 0.615 | | | | Accuracy: 61.9%；F1-score: 0.528 | | | |

**Supplemental Table 5. Identification of tissue-of-origin by coverage profile for cancer patients in 5hmC sequencing data (Feature number: 39)**

| Cancer | Validation set | | | | Test set | | | |
|---|---|---|---|---|---|---|---|---|
| type | Precision | Recall | F1-socre | Patients | Precision | Recall | F1-socre | Patients |
| GBM | 0.76 | 0.93 | 0.84 | 15 | 0.67 | 0.71 | 0.69 | 14 |
| HCC | 0.85 | 0.88 | 0.87 | 26 | 0.81 | 0.93 | 0.86 | 27 |
| LUAD | 0.67 | 0.29 | 0.40 | 7 | 0.67 | 0.29 | 0.40 | 7 |
| PDAC | 0.73 | 0.73 | 0.73 | 15 | 0.71 | 0.67 | 0.69 | 15 |
| All types | Accuracy：79.0%；F1-socre: 0.710 | | | | Accuracy: 74.6%；F1-score: 0.660 | | | |

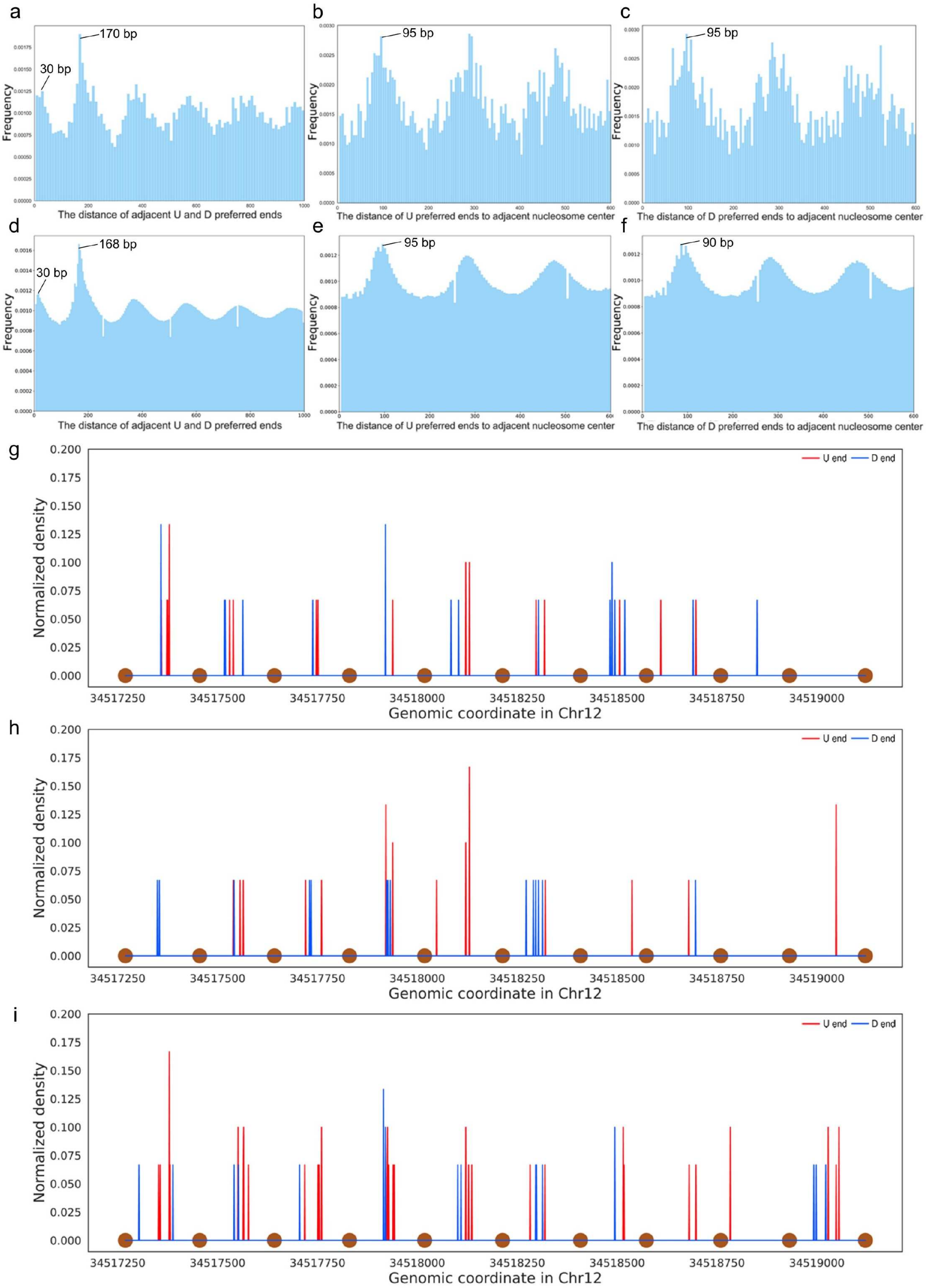
a
170 bp
30 bp
Frequency
The distance of adjacent U and D preferred ends
b
95 bp
Frequency
The distance of U preferred ends to adjacent nucleosome center
c
95 bp
Frequency
The distance of D preferred ends to adjacent nucleosome center
d
168 bp
30 bp
Frequency
The distance of adjacent U and D preferred ends
e
95 bp
Frequency
The distance of U preferred ends to adjacent nucleosome center
f
90 bp
Frequency
The distance of D preferred ends to adjacent nucleosome center
g
U end
D end
Normalized density
0.200
0.175
0.150
0.125
0.100
0.075
0.050
0.025
0.000
34517250
34517500
34517750
34518000
34518250
34518500
34518750
34519000
Genomic coordinate in Chr12
h
U end
D end
Normalized density
Genomic coordinate in Chr12
i
U end
D end
Normalized density
Genomic coordinate in Chr12

Supplementary Figure 1. The distribution of preferred ends related to the position of the nucleosomes.
(a-c) Histgram showing distance distribuition between adjcent U and D ends, and the nucleosome center in a regions that is known to be positioned nucleosomes in almost all non-malignant tissue types (chr12: 34376000-34452000) [11,12]. The distance between any U to its nearest D ends reached the first peak at ~30bp, which was about the size of a DNA linker (20-40bp) [13]. The length of the DNA stretch that wrapped around nucleosomes was reported to be 146bp [14], which was exactly the distance between the first and second nearest D end to any U end (the first and second peak, a).
The distance between D/U ends and nucleosome centers was ~80 to 100bp, which was half of the size of one nucleosome wrapped DNA stretch plus two DNA linkers (b-c). (d-f) Histgram showing distance distribution of adjacent U and D preferred ends, and the nucleosome center in chromosome 12. (g-h) Distribuition of cfDNA preferred end signals in a nucleosome array region (chr12: 34517269-34519122) in GBM (g), LUAD (h) and PDAC (i) subjects (n=15 in every cancer type). Brown dots at the bottom represent the predicted nucleosome center loci reported in a previous study [11,12].

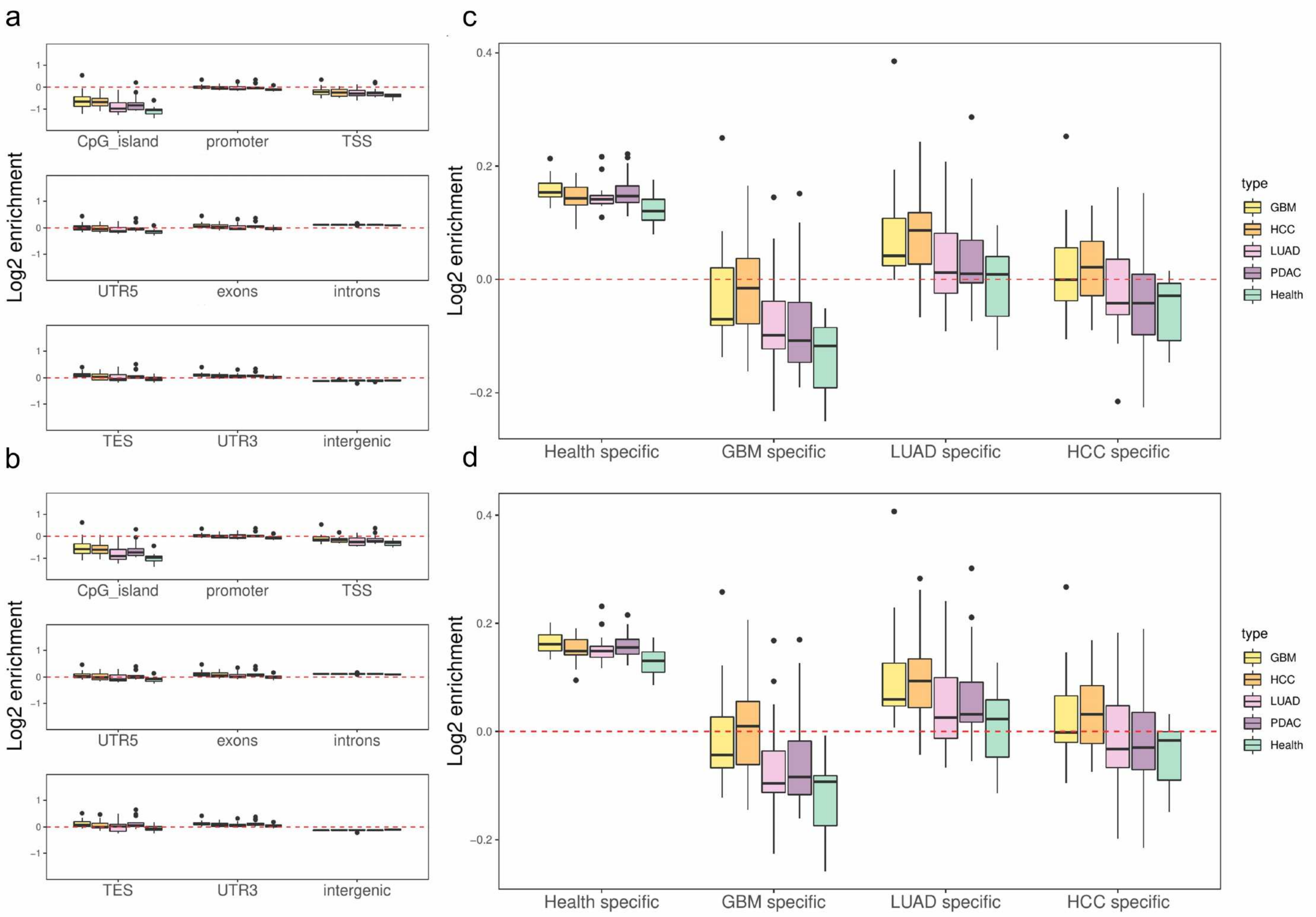


Supplementary Figure 2. Enrichment of preferred ends found in WGS data in genomic features and open chromatin regions.

Boxplot showing Log2 enrichment of U (a) and D (b) preferred ends in genomic features, and U (c) and D (d) preferred ends enrichment in open chromatin regions.

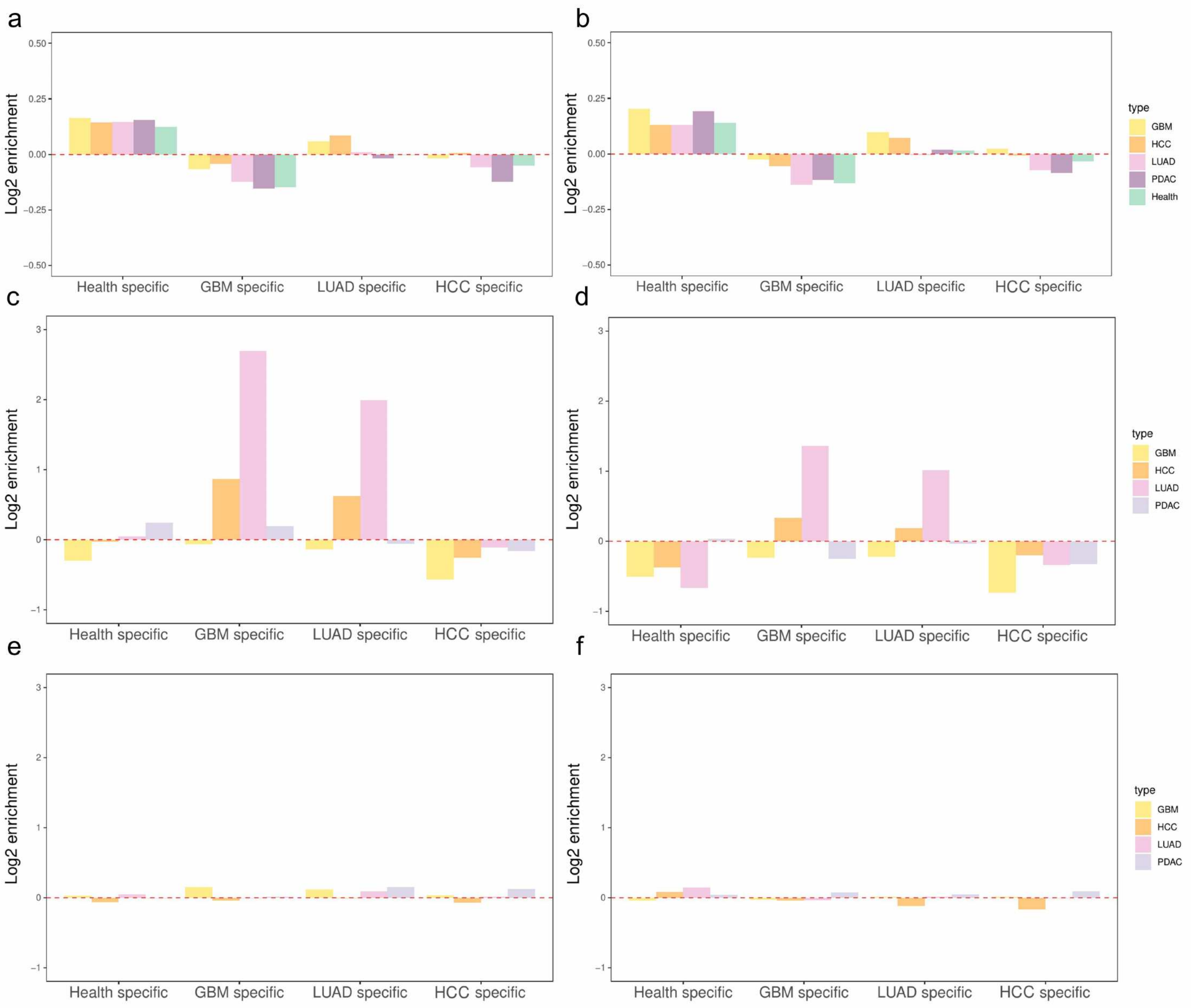


Supplementary Figure 3. Enrichment of preferred ends found in WGS data in open chromatin regions.

Barplot showing Log2 enrichment of repeatable U (a) and D (b) preferred ends, cancer-specific U (c) and D (d) preferred ends and health-specific U (e) and D (f) preferred ends in open chromatin regions. Repeatable preferred end: the preferred end appeared in at least two samples in each cancer type.

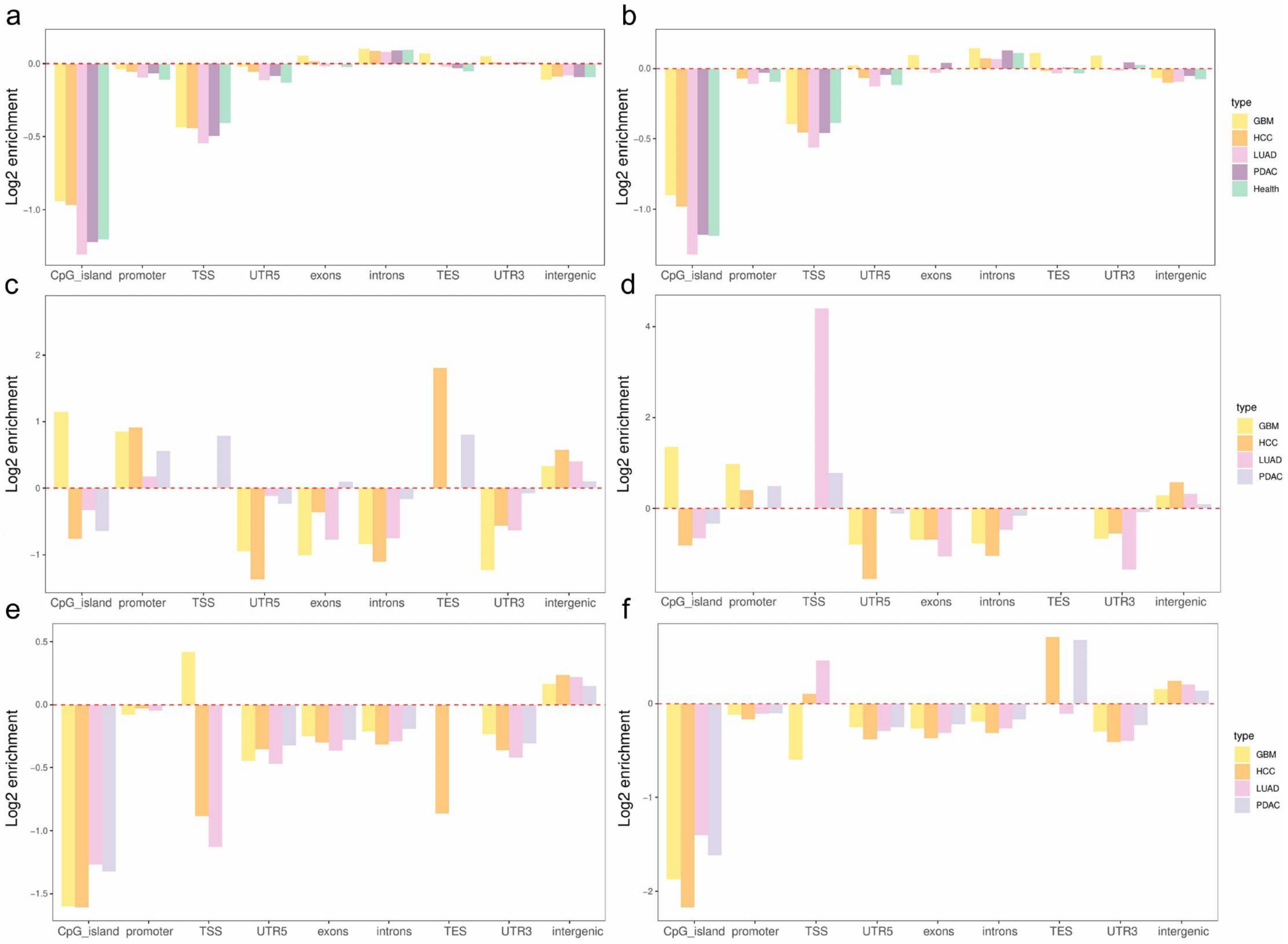


Supplementary Figure 4. Enrichment of preferred ends found in WGS data in genomic features.
Barplot showing Log2 enrichment of repeatable U (a) and D (b) preferred ends, cancer-specific U (c) and D (d) preferred ends and health-specific U (e) and D (f) preferred ends in genomic features.

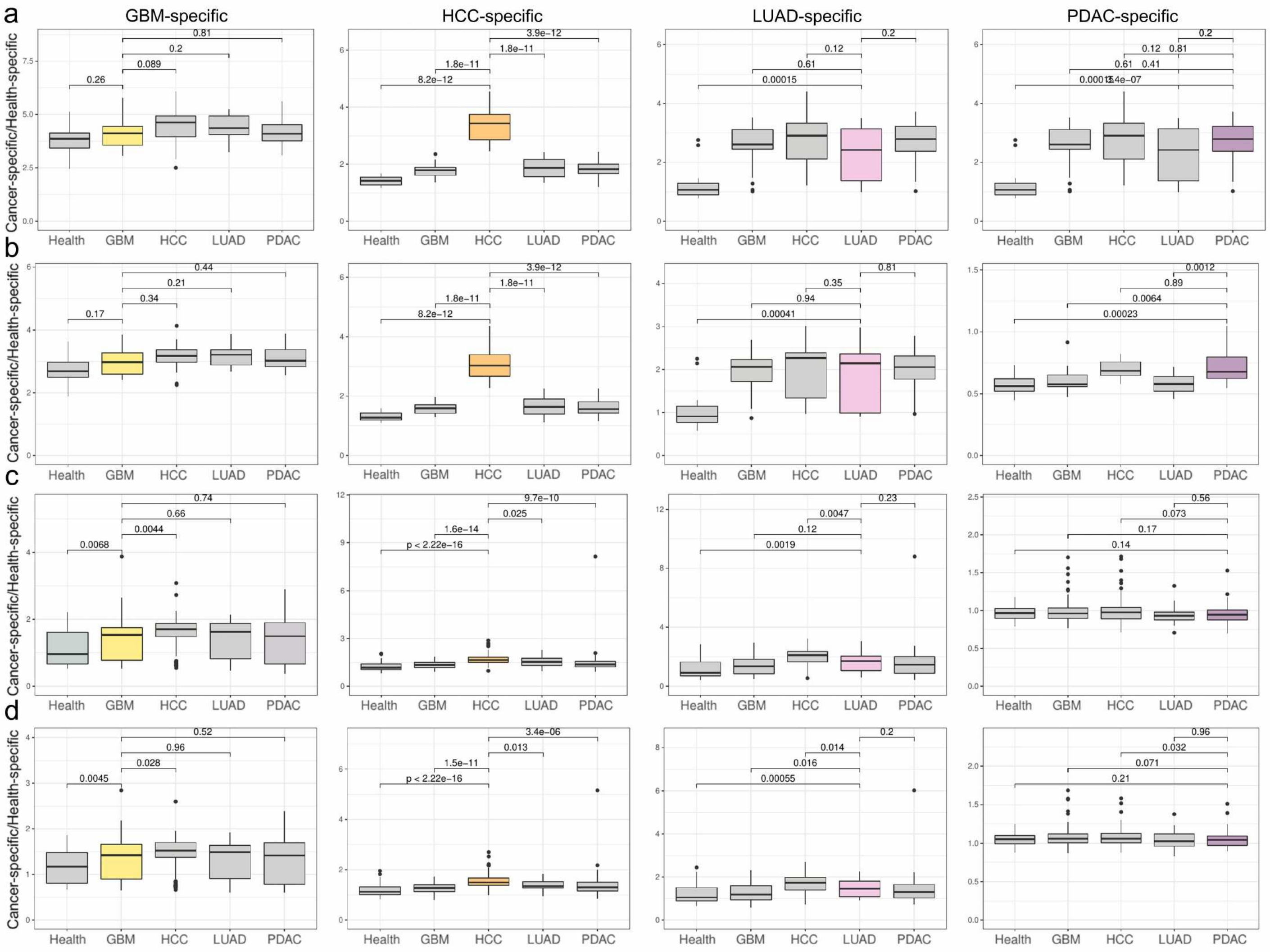


Supplementary Figure 5. Preferrence of preferred ends found in WGS data regard to sample types.

(a-b) Boxplots showing the ratios of the cancer specific to health specific preferred U (a) and D (b) ends in low-pass WGS samples (Wilcoxon rank-sum test). (c-d) Boxplots showing the ratios of the cancer specific to health specific preferred U (c) and D (d) ends in 5hmC sequencing data (Wilcoxon rank-sum test).

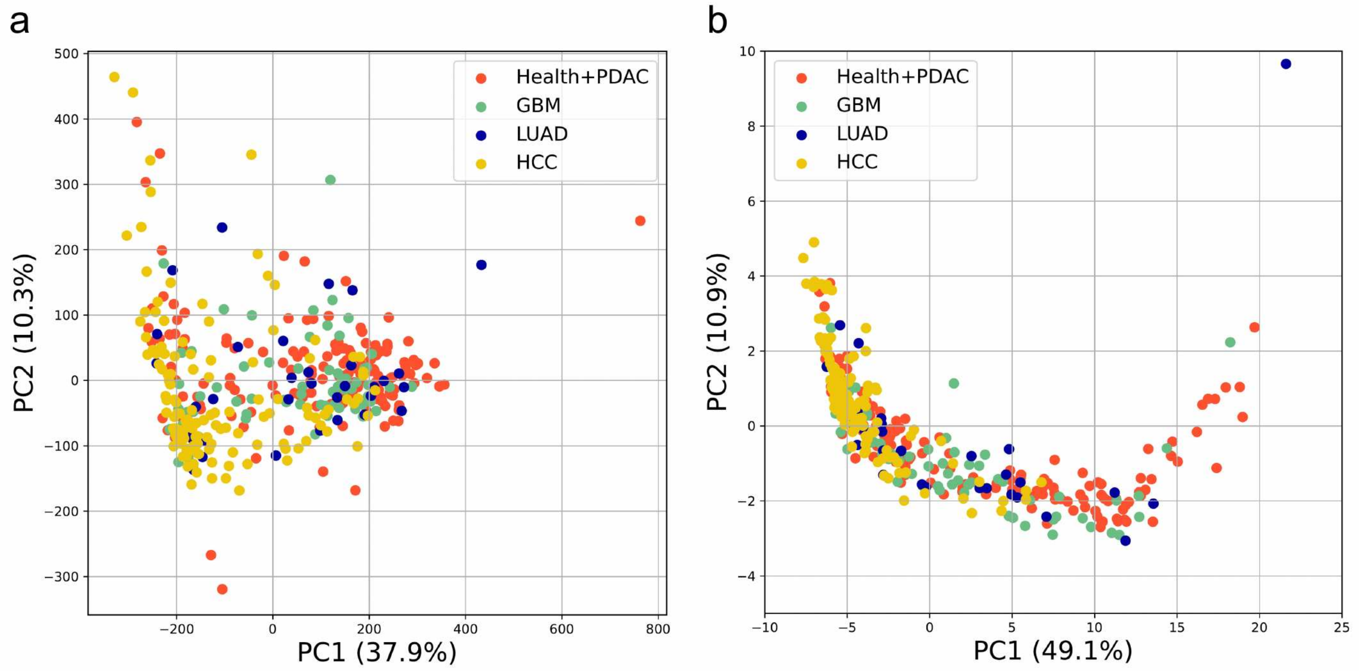


Supplementary Figure 6. PCA plots showing clustering of 5hmC sequencing samples based on (a) coverage profile and (b) size profile.

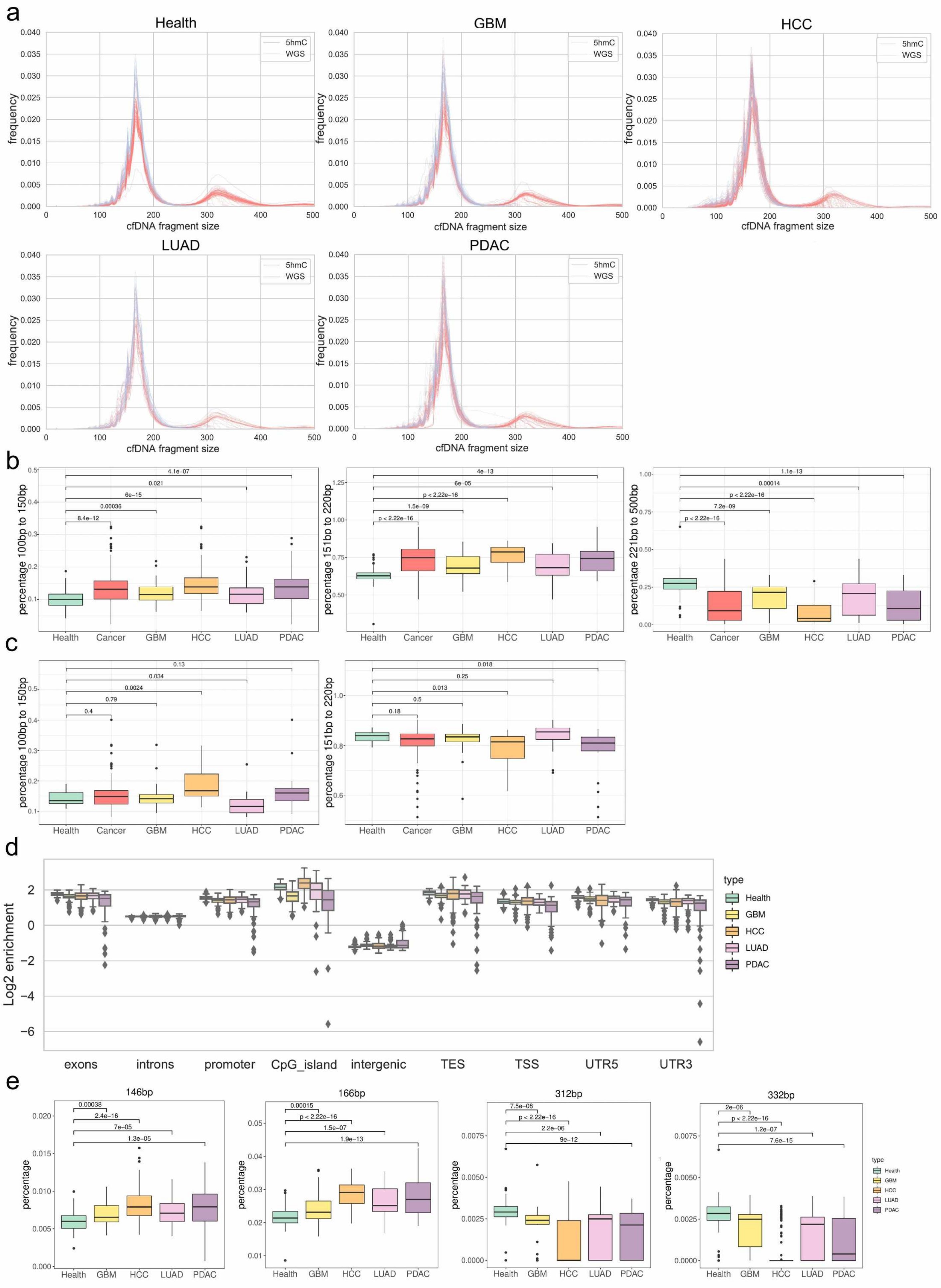
a
Health
GBM
HCC
LUAD
PDAC
5hmC
WGS
frequency
cfDNA fragment size
b
percentage 100bp to 150bp
percentage 151bp to 220bp
percentage 221bp to 500bp
4.1e-07
0.021
6e-15
0.00036
8.4e-12
4e-13
6e-05
p < 2.22e-16
1.5e-09
1.1e-13
0.00014
7.2e-09
Health
Cancer
GBM
HCC
LUAD
PDAC
c
0.13
0.034
0.0024
0.79
0.4
0.018
0.25
0.013
0.5
0.18
d
Log2 enrichment
type
exons
introns
promoter
CpG_island
intergenic
TES
TSS
UTR5
UTR3
e
146bp
166bp
312bp
332bp
percentage
0.00038
2.4e-16
7e-05
1.3e-05
0.00015
1.5e-07
1.9e-13
7.5e-08
2.2e-06
9e-12
2e-06
1.2e-07
7.6e-15

Supplementary Figure 7. cfDNA size profile of 5hmC sequencing data and WGS data.

(a) Comparison of frequency distribuition of cfDNA size between 0-500bp between WGS data and 5hmC sequencing data. (b) Boxplots showing percentage of short (100-150 bp) fragments, long (151-220 bp) fragments and ultra-long fragments (221-500 bp) in 5hmC sequencing data (Wilcoxon rank-sum test). (c) Boxplots showing percentage of short (100-150 bp) fragments, and long (151-220 bp) fragments in WGS data. (d) Enrichment of ultra-long fragments with above 1 5hmC peak in genomic features. (e) cfDNA fragments percentage in size of one nucleosome (146 bp), one nucleosome plus one DNA linker (166 bp), two nucleosomes plus one DNA linker (312 bp), two nucleosomes plus two DNA linkers (332 bp) in 5hmC sequencing data (Wilcoxon rank-sum test).

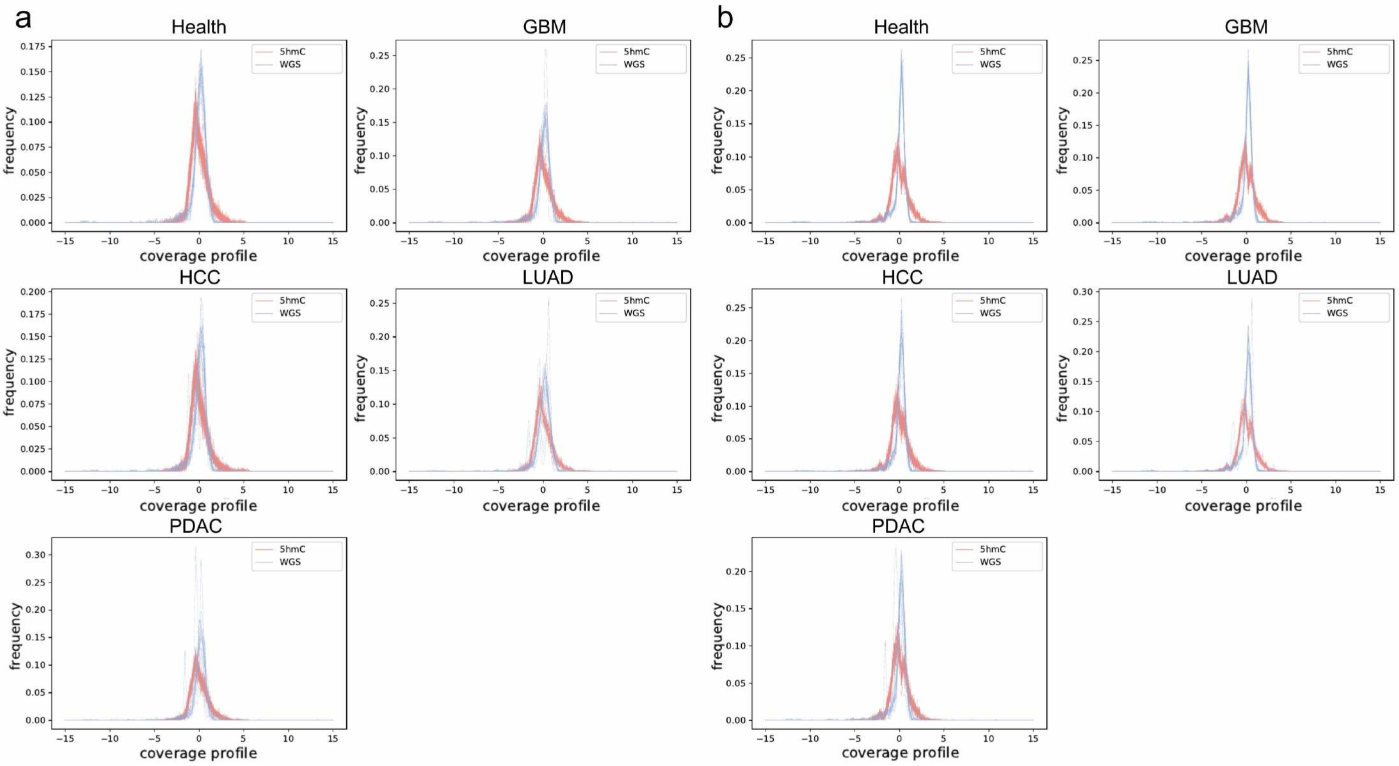


Supplementary Figure 8. Comparison of coverage profile of (a) short cfDNA fragments and (b) long cfDNA fragments between low-pass WGS data and 5hmC sequencing data. Mean value of the percentage of coverage profile between -1 and 1 was 95% in WGS data and 86% in 5hmC data for short fragments, and 98% in WGS data and 86% in 5hmC data for long fragments, Student's t-test, p value < 0.001.

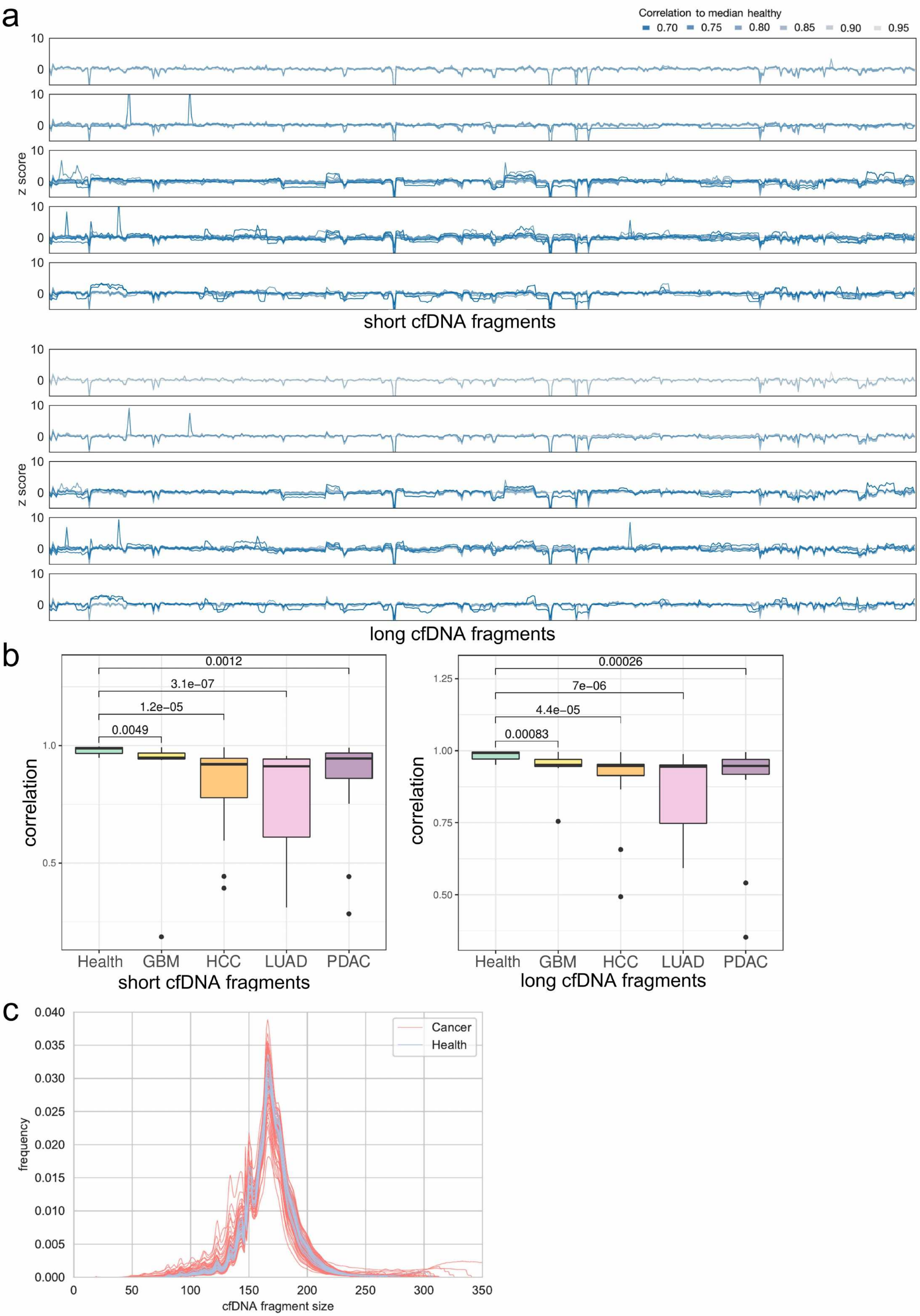
a
Correlation to median healthy
0.70 0.75 0.80 0.85 0.90 0.95
z score
10
0
short cfDNA fragments
long cfDNA fragments
b
correlation
0.0012
3.1e−07
1.2e−05
0.0049
1.0
0.5
Health GBM HCC LUAD PDAC
short cfDNA fragments
0.00026
7e−06
4.4e−05
0.00083
1.25
1.00
0.75
0.50
long cfDNA fragments
c
frequency
0.040 0.035 0.030 0.025 0.020 0.015 0.010 0.005 0.000
Cancer
Health
0 50 100 150 200 250 300 350
cfDNA fragment size

Supplementary Figure 9. Evaluation of cfDNA size profile and coverage profile in WGS data.

(a) Genome-wide coverage profile of short and long cfDNA fragments, color indicated sample-wise correlation to median health. (b) Boxplots showing corrleation to median healthy in health cohort and 4 types of cancer cohort in short and long cfNDA coverage profile (Wilcoxon rank-sum test). (c) Comparison of cfDNA size profile of WGS data between cancer and controls. Mean value of the standard deviation of density for healthy controls was 0.00032, while 0.00070 for cancer patients in the range of 1-300bp.

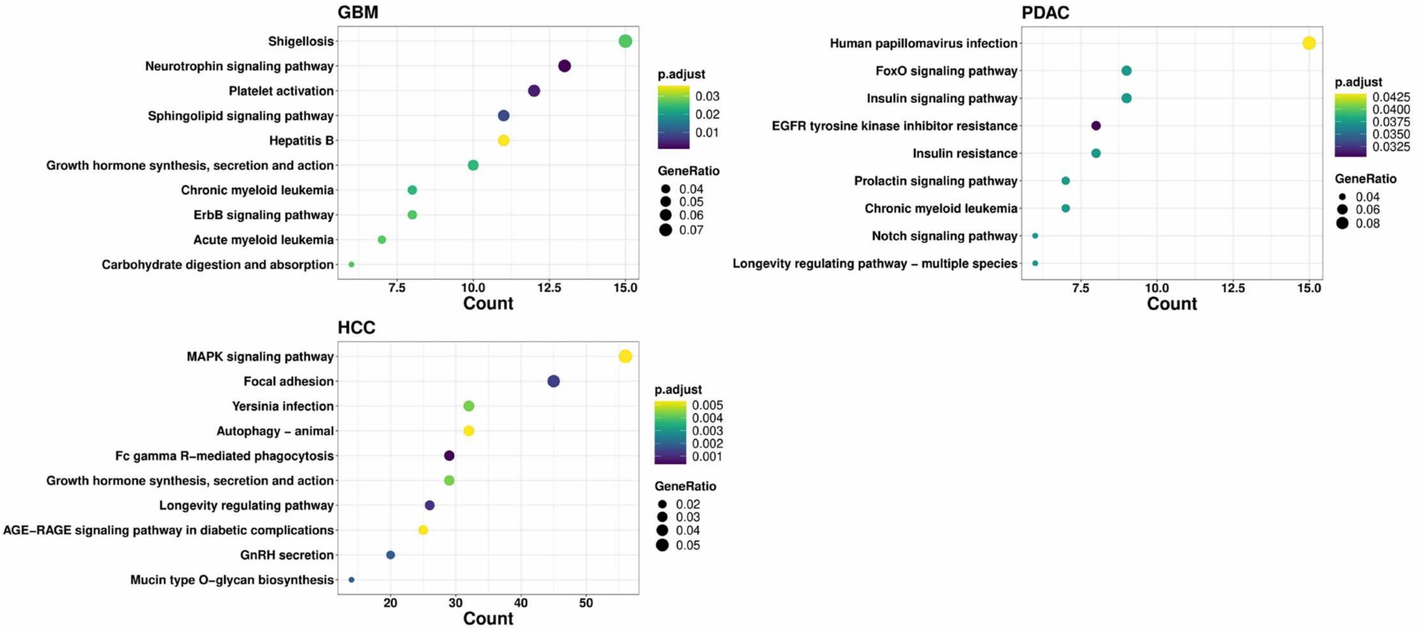


Supplementary Figure 10. KEGG enrichment result of genes located within differentially modified 5hmC peaks detected in GBM, HCC and PDAC cohort.

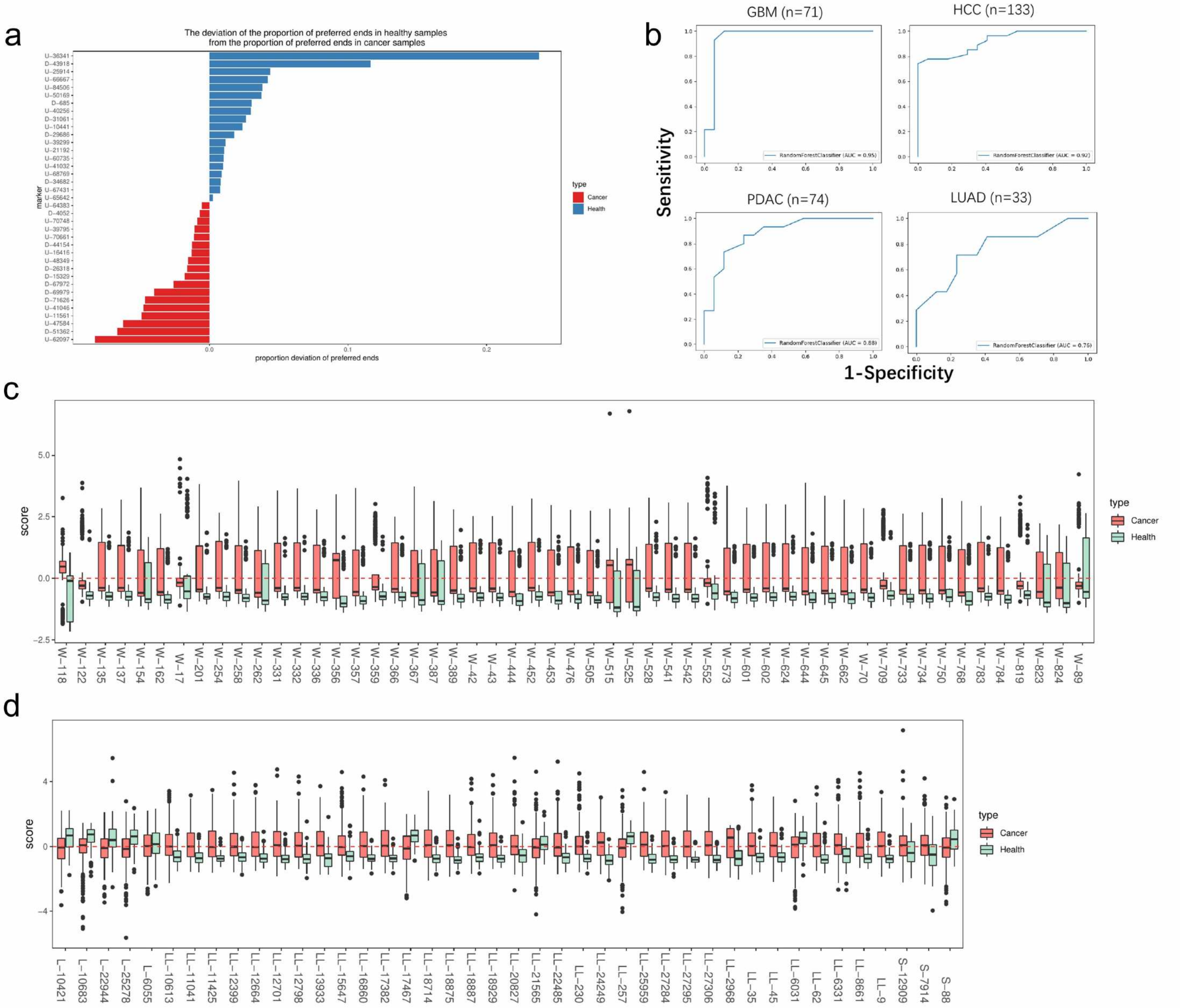


Supplementary Figure 11. cfDNA fragmentomic features for pan-cancer diagnosis using 5hmC sequencing data.

(a) The deveiation of proportion of preferred ends in healthy samples from the proportion of cancer samples. (b) ROC curves for the integrated model in single cancer detection. (c) Performance of the 5hmC signature biomarkers in the 5hmC signature alone model. (d) Performance of the coverage profile biomarkers in the coverage profile alone model.
